# Weight loss in patients with COVID-19 and Influenza in comorbidity with NCDs: a pilot prospective clinical trial

**DOI:** 10.1101/2022.12.04.22283077

**Authors:** Kuat Oshakbayev, Aigul Durmanova, Alisher Idrisov, Zulfiya Zhankalova, Gulnara Bedelbayeva, Meruyert Gazaliyeva, Attila Tordai, Altay Nabiyev, Bibazhar Dukenbayeva

**Author notes:** **Corresponding Author:** Kuat Oshakbayev. University medical center, Internal Medicine Department. The Republic of Kazakhstan, 010000, Astana, str. Syganak, 2. .mob.

## Abstract

**Background:** COVID and Influenza with non-communicable chronic diseases (NCDs) complicate the diagnosis, treatment, prognosis, and increase mortality rate. The aim: to evaluate the effects of the fast weight loss on clinic and laboratory inflammation profile, metabolic profile, reactive oxygen species (ROS) and body composition in patients with COVID and Influenza in comorbidity with NCDs.

**Methods:** A 6-week open, pilot prospective clinical trial including 62 adult patients with COVID (n=27) and influenza (n=35) in comorbidity with T2D, hypertension, and NASH. Overweight in 33 patients (53.2%) with BMI 28.14±0.39 kg/m^2^, and 29 patients without overweight with BMI 23.37 ± 0.38 kg/m^2^. T2D in 26 (41.9%); Hypertension in 38 (61.3%) (incl. 12 patients with T2D); NASH in 51 patients (82.2%) (incl. 8 patients with NASH, T2D and Hypertension; 6 patients with NASH and T2D; 18 patients with NASH and Hypertension; 19 patients with only NASH). Primary endpoints: Clinic/infectious/inflammation tests for COVID and Influenza; weight loss during 14 days. Secondary endpoints: fasting blood glucose, HbA1c, blood insulin; systolic/diastolic BP; blood lipids; ALT, AST, chest CT-scan.

**Results:** The patients with overweight lost -12,4% from baseline or BMI= -4.2 kg/m^2^, and patients without overweight lost -9,14% from baseline or BMI= -2.2 kg/m^2^ (−9.7±0.7 kg vs. - 6.4±0.6 kg, respectively; *P*<0.001) at 14-day of the treatment. Weight loss in both groups was due to reduction of fat mass (*P*<0.0001).

Sputum production increased in 1.0-1.5 liter/day on 2-3 days, decreased in 7-9 days. Body temperature normalized in 6-9 days. On 3-5 days, in most patients their urine became turbid/muddy/intensively colored. Urine microscopy showed organic and non-organic salts, and leukocyturia (20-35/sight).

White blood cells, lymphocytes, NLR normalized at 14 days (P<0.0001). Total-fibrinogen, C-reactive-protein, and Erythrocyte-sedimentation-rate, ROS normalized at 14-day of treatment (*P*<0.0001).

COVID and Influenza were a negative in >96.3% patients at 14-day. Systolic/diastolic BP decreased (161.3±1.31/101.6±0.85 vs. 118.3±0.46/80.89±0.66, *P*<0.0001), glucose and lipids metabolism in patients with T2D (n=26) (P<0.0001); ALT and AST in patients with NASH (n=51) were significantly normalized (from baseline 134.3±5.4 and 166.5±5.5 U/L, respectively, and at 14-day to 78.4±4.2 and 92.4±4.9 U/L, respectively (P<0.0001)), platelets increased from baseline (186.5±4.6, ×10^9^/L) at 14-day of treatment (238.5±5.8, ×10^9^/L) (*P*<0.0001), and at 6-week follow-up (278.3±6.9, ×10^9^/L) (*P*<0.0001). The mean score of chest-CT for the patients (n=44) was 13.12±0.38 from baseline, and at 14-day the score was 1.72±0.12 (*P*<0.0001). ROS level normalized at 14-day treatment and 6-week follow-up from baseline (*P*<0.0001). The previous antidiabetic, antihypertensive, anti-inflammatory and hepatoprotective, and other symptomatic medications were adequately decreased in 2-5 days to completely stopping by 5-8 days treatment.

**Conclusions:** The fast weight loss is clinical/laboratory benefit in treatment of patients with COVID-19 and Influenza in comorbidity with T2D, hypertension, and NASH.

**Trial Registration:** ClinicalTrials.gov NCT05635539. Registered 1 December 2022.

## 1. INTRODUCTION

SARS-CoV-2, a virus which causes the disease known as COVID-19 (further – COVID), was first described in a case of pneumonia of unknown origin in Wuhan City, China but quickly evolved into a worldwide pandemic, and has changed the mortality rate. (1-3) COVID and Influenza are the most common acute respiratory diseases (ARD) in the world, which can quickly spread through the air and direct contact. (2, 4, 5) The ARDs are serious illnesses that >100 millions of people in the worldwide get each year. It sends millions of people to the hospital and causes millions of deaths. (6, 7) The increased ranking of COVID as a leading cause of death in some age groups is consistent with a downward age shift in the distribution of COVID deaths in last years. (3, 8)

COVID and Influenza co-infection with other non-communicable chronic diseases (NCDs) complicate the diagnosis, treatment, prognosis of the ARDs emerge new concern. (9, 10) This comorbidity may even increase the disease symptom and mortality rate. Aggravating affects of the ARDs on cardiovascular comorbidities, diabetes onset, and chronic liver and kidney diseases have long been debating. (11, 12) Older age and type 2 diabetes (T2D), hypertension, liver and kidney diseases were significantly associated with an increased likelihood of mortality. (13) NCDs are also leading cause of mortality and morbidity in the world. (14, 15) Pharmacology treatment of ARDs and NCDs evidenced positive results, but it is still insufficiently, due to many problems related with their effectiveness, wide side-effects, sensitivity and specificity, etc. (14, 16, 17) Vaccination for controlling COVID and Influenza infectious diseases continues to be the most effective method. Nonetheless, COVID and Influenza variants continue to evolve and emerge, resulting in significant public concerns worldwide. (3, 18)

Pharmacologic and bio-vaccine treatment are not always can help to cure the ARDs due to they have a lot of side effects during and post treatment period. Treatment of the ARDs is more difficult in patients with comorbidities as T2D, hypertension, and nonalcoholic steatohepatitis (NASH). (19-21) Therefore, searching reliable, secure, and natural methods for treatment of COVID and Influenza are continuing in the worldwide. (3, 5, 17, 18, 22)

NCDs and COVID are spreading in countries with a higher rate of overweight. (10, 23-27) The Comprehensive Assessment of the Long-term Effects of Reducing Intake of Energy (CALERIE) phase 2 trial showed that moderate reduction of energy intake averaging approximately 12% over 2 years could improve markers of inflammation, cardiometabolic health, and oxidative stress in humans. (28, 29) Caloric restriction has been shown to extend life span and health span in many animal species.(30) There has been growing interest in evaluating related strategies, such as intermittent fasting, periodic fasting, and time-restricted eating that may achieve the putative benefits of caloric restriction with greater likelihood of sustainability. (31) COVID affects different people in different ways. Most infected people will develop mild to moderate illness and recover without hospitalization. Some studies reported a high prevalence of overweight and obesity in patients experiencing a severe COVID course, with serious complications requiring hospitalization and admission to intensive care units. (27) Patients with obesity require regularly taking medications for the treatment of any concurrent chronic diseases, their physicians must promptly manage any medical symptoms in the case of suspected acute respiratory syndrome infection to prevent the risk of severe outcomes. (32) ARDs with overweight are associated with an increase in oxidative stress and a decrease in antioxidant protection. (23) Acute inflammation in patients with overweight extreme difficulties in treatment and highlight the need for preventive measures. (33)

Pharmacological treatment of ARDs with NASH is a great challenge for medicine, because the patients are often limited to take medicine due to persistent progression, increasing of hepatocellular injury/ inflammation, and medicinal overloading. (20)

In previous studies we showed a significant impact of overweight/obesity prevalence on the increase in COVID morbidity/mortality, (26) and beneficial role of our weight loss method in improving glycemic, lipid and hormone profiles, electrolyte and biochemical indices, blood pressure, reactive oxygen species and bone mineral density in patients with T2D, hypertension, and severe NASH. (34, 35) The aim of the study was: to evaluate the effects of our fast weight loss method on clinic and laboratory inflammation profile, metabolic profile, reactive oxygen species (ROS) level and body composition in patients with COVID and Influenza in comorbidity with T2D, hypertension, and NASH.

## 2 STUDY DESIGN AND PARTICIPANTS

### Study Design

A 6-week, open, pilot prospective clinical trial with the intention-to-treat principle.

#### 2.1 Participants

The study enrolled 72 adult people (38 women) aged from 25 to 80 years with moderate-to-severe cases COVID and Influenza in comorbidity with NCDs as T2D, hypertension, and NASH; 6 patients were excluded due to noncompliance of inclusion and exclusion criteria. Of the remaining 66 patients 4 patients (6.5%) dropped out prior to the study completion: 2 patients refused the treatment method in 3 days after starting, and 1 patient moved to another city, 1 patient was excluded due to noncompliance. (**Fig. 1**) A control pharmacology group was not used because medicinal treatment was unacceptable due to NASH in most of the patients. Thus, 62 patients (36 women) included for the analysis; 27 patients with COVID and 35 patients with Influenza. Overweight in 33 patients (53.2%) with body mass index (BMI) 28.14±0.39 kg/m^2^, and 29 patients without overweight with BMI 23.37 ± 0.38 kg/m^2^. T2D in 26 (41.9%); Hypertension in 38 (61.3%) (incl. 12 patients with T2D); NASH in 51 patients (82.2%) (incl. 8 patients with NASH, T2D and Hypertension; 6 patients with NASH and T2D; 18 patients with NASH and Hypertension; 19 patients with only NASH). Thus, all patients with the ARDs had in comorbidity with one or more NCDs. All patients with T2D were on Metformin, Sulfonylurea, DPP-4 inhibitors, GLP-1 receptor agonists, Glitazones in different combinations. All patients with hypertension received antihypertensive treatment. All patients refused for pharmacology therapy due to: either previous unsuccessful drug results; or an antimicrobial resistance profile; or drug allergy; or reluctance to take medication; or iatrogenic fear (iatrophobia); or a rich failed experience in drug treatment; and NASH.

**Figure 1.**
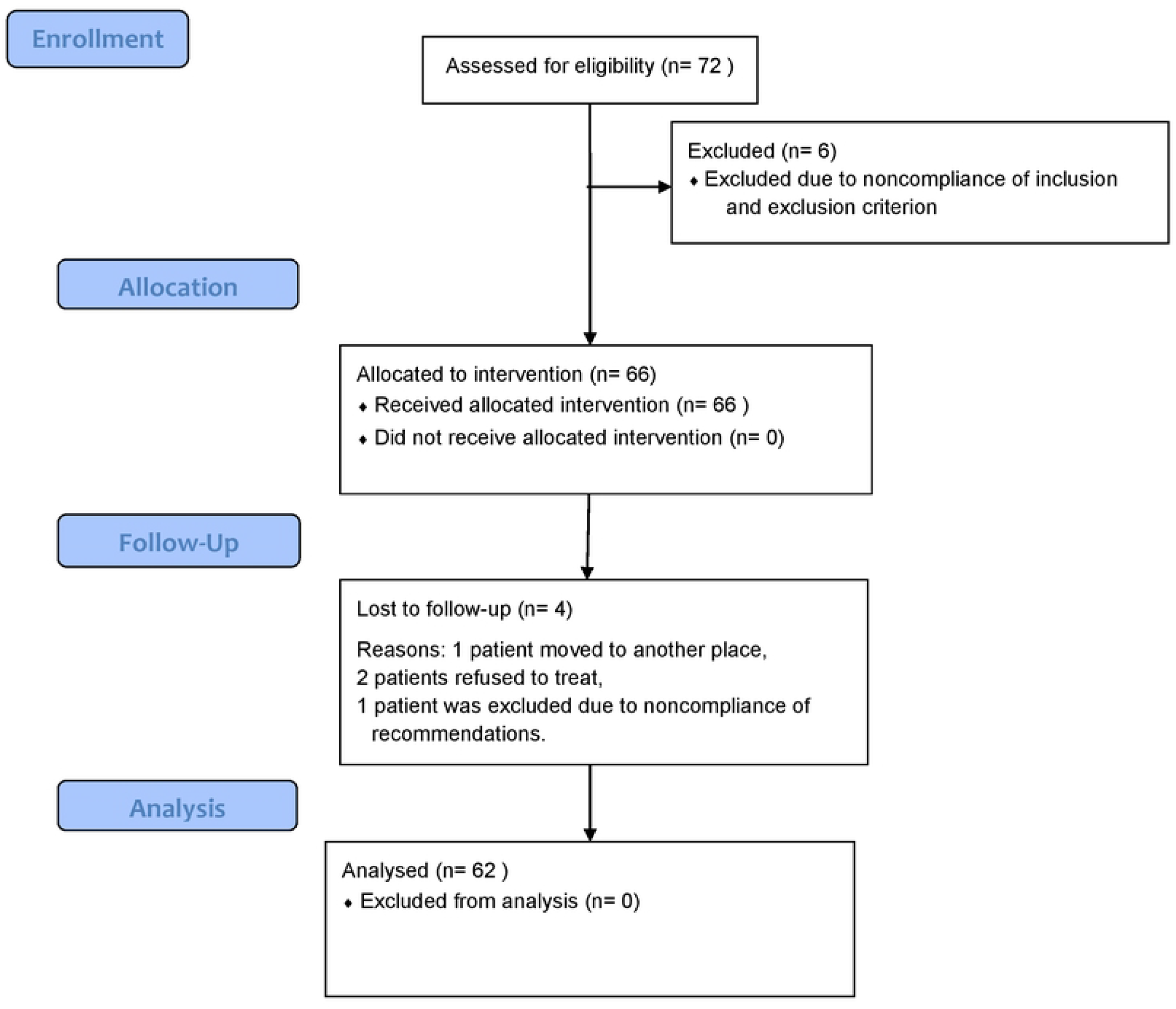
Participant Flow Diagram

All the patients were admitted into the out-patient department in 3-5 days after illness onset. The participants were recruited gradually as they come in the Republican Diagnostic Center at University Medical Center (Astana) and ANADETO medical center from November 2020 to March 2022. The study was carried out in the Republic of Kazakhstan from November, 2020, through July, 2022.

##### Inclusion criteria

1) written informed consent form; 2) patients with fever; 3) patients refused for pharmacology therapy; 4) weight loss treatment for 12-14 days and +4 weeks follow-up (total 6 weeks).

##### Exclusion criteria

1) patients with acute respiratory failure and assisted ventilation requirement; 2) respiratory rate ≥ 30 times per minute; 3) oxygen saturation ≤ 93% by finger oximetry at resting status.

##### Outcome measures

*Primary endpoints:* Clinic and infectious tests for COVID and Influenza; weight loss during 14 days; inflammation profile. *Secondary endpoints:* fasting blood glucose, glycosylated hemoglobin A1c (HbA1c), blood insulin; systolic/diastolic blood pressures (BP); blood lipids; alanine aminotransferase (ALT), aspartate aminotransferase (AST), chest computed tomography (CT) scan.

#### 2.2 Analytical Assessment

##### Pathogen detection methods

###### Infection detection

COVID was diagnosed primarily by direct detection of SARS-CoV-2 RNA by nucleic acid amplification tests with a real-time reverse-transcriptase-polymerase-chain-reaction (RT-PCR) assay using nasal or pharyngeal swab specimens from the upper respiratory tract. A health care professional collected a fluid sample by inserting a long nasopharyngeal swab into a nostril and taking fluid from the back of a nose.

To diagnosis of the flu was used a RT-PCR test called the Flu SC2 Multiplex Assay. Sputum DNA was extracted using Chemagic DNA kits (PerkinElmer, Turku, Finland) and PCR assays were performed using Seeplex Pneumobacter ACE Detection kits (Seegene Inc., Seoul, South Korea) according to the manufacturer protocol. (36)

Sputum specimens collection, samples transport, samples storage, diagnostic test procedures environment, to tests results reporting, every step were strictly performed under the standard guidelines. (37, 38)

All the tests were performed in two independent laboratories in Astana: BioLab laboratory and OLIMP laboratory. Positive results were considered when both laboratories give equal results. *Biochemical laboratory assessments* consisted of a complete blood cell count, white blood cells (normal: 3.5-9.0×10^9^/L), neutrophil-to-lymphocyte ratio (NLR) is calculated by dividing the absolute count for neutrophils by the absolute count for lymphocytes (normal: 1-2), blood chemical analysis, erythrocyte sedimentation rate, urea (normal: 2.1-7.7 mmol/L), creatinine (normal: 53-115 μmol/L), glucose, cholesterol (normal: <5.4 mmol/L), triglyceride (normal: <1.69 mmol/L), high-density lipoprotein (HDL) (normal: >1.55 mmol/L), fibrinogen (normal: 2.0-4.0 g/L), hepatic enzyme levels (ALT, norm <36 U/L, AST, norm <40 U/L), assessment of liver and renal function, and measures of inflammation level, C-reactive protein (normal: ≤ 10.0 mg/L), and HbA1c (normal: <5.7%; prediabetes: 5.7-6.5%, diabetes: ≥ 6.5%). All the data of enrolled cases, including demographic information, clinical symptoms or signs, clinical outcomes and laboratory findings on admission, were included in electronic medical records.

###### Hormonal assays

Fasting serum insulin was determined by immunoassay (Immunotech Insulin Irma kit, Prague, Czech Republic). Hyperinsulinemia was considered >12.5 nU/L. The Homeostasis Model Assessment insulin resistance indexes (HOMA-IR) was used as surrogate measure of insulin sensitivity as follows: HOMA-IR = ((fasting insulin in nU/L) × (fasting glucose in mmol/l)/22.5). Insulin resistance was considered if the index was >2.

Diagnostic criteria for T2D were used the standards of American Diabetes Association in 2017. (39) Hypertension was diagnosed by BP readings and from medical records. Physical activity of patients was assessed by the number of steps measured by pedometers (Hoffmann-La Roche Ltd, Basel, Switzerland).

The Nonalcoholic Steatohepatitis Clinical Research Network criteria was used to diagnose NASH and from medical records. (40)

###### Anthropometrical data

Anthropometrical indicators included age (years), weight (kg), BMI (kg/m^2^). Body composition parameters including fat mass (in % of total body weight and total kg), fat free mass (kg), total body water (%), muscle mass (%), and bone mass (%) were measured using a Tanita-SC330S Body Composition Analyzer (Tanita Corp., Tokyo, Japan). *For ROS we measured* malondialdehyde (MDA, a marker of lipid peroxidation and degradation product of oxidized fatty acids), superoxide dismutase (SOD) and catalase (antioxidant enzymes), and advanced oxidation protein products (AOPP, markers of protein oxidation). MDA (μmol/L) was measured using a thiobarbituric acid-reactive substance assay as described by Yagi and modified by Ohkawa. (41)

Antioxidant enzymes were determined in erythrocyte hemolysates, in which hemoglobin concentration was assayed using Drabkin’s method. Superoxide dismutase was measured by the method of Misra and Fridrovich. (42) Catalase was measured at λ =240 nm at 25 °C; catalase activity was expressed in U/g and one unit is considered as the amount of the enzyme which decomposes 1 g of H_2_O_2_ in 1 min at 25 °C in pH =7.0. AOPP (μmol/L) were measured by adding 40 μL of plasma to a 190 μL of mixture of 81% phosphate buffer solution, 10% glacial acetic acid and 4% 1.16 mmol solution of KI, followed by 2 minutes absorbance at 340 nm on a plate reader Multiscan Ascent using a spectrophotometric method. Chloramine-T solution was used as calibrator. (43)

###### Imaging

Ultrasound images (GE Vivid 9 Ultrasound; GE Healthcare Worldwide USA, Michigan) and chest CT (Siemens Somatom Sensation 32) were obtained. The key imaging finding in the pneumonia were ground-glass opacity, distribution and extent of lung abnormalities, including consolidation, cavitation and pleural effusion.

Chest CT assessment of the severity of lung changes. Each lung field was divided into three equal zones. Each zone was assigned a score from 0–4 based on the percentage of lung involved (0 = no abnormality, 1 = <25% of the zone involved, 2 = 25–50% involved, 3 = 51– 75% involved and 4 = >75% involved). The scores for all six zones of each CT examination were summed to provide a cumulative chest radiographic score (range, 0–24). (44, 45) Baseline and follow-up CT scans were reviewed in consensus by two radiologists with 10–25 years of experience. Laboratory tests, ultrasound and CT images, and an electrocardiogram were performed, and sputum and blood samples were collected on the inclusion day.

###### Intervention

We used the “Analimentary detoxication” (ANADETO) weight loss method, including calorie restriction to 50-100 kcal/day with fat-free vegetables (tomatoes and cucumbers) with mandatory salt intake to 5-6 gr/day, hot water drinking 1000-1500 ml/day, walking at least 2,000 steps/day after normalized body temperature, and sexual self-restraint. The walking provided to promote of blood circulation and decrease in metabolic intoxication. The weight loss method lasted 14 days. (46-48) Then the patients followed for 4-week a diet where they ate one meal a day without any food restriction. A combination of in-person conversations and telephone calls were conducted during whole 6-week study period.

###### Ethics

The Local Ethics Committee of the University Medical Center (phone: +71272-692586; https://umc.org.kz/en/?ethics-commission) approved the study “The fast weight loss in patients with acute and chronic respiratory infections in comorbidity with non-communicable diseases” (approval protocol number is #7 of 26.09.2019).

### Statistics

The two-side Student’s t-test and Odds ratios (ORs) with 95% Confidence intervals (CIs) were used. The study data were tested against the normal distribution and are presented in Tables as Mean±Standard Error of the Mean (M±SEM). *P*-value of <0.05 was set as significant and <0.0001 was set as highly significant. Statistical analysis was performed using SPSS ver.23.0 for Windows (SPSS: An IBM Company, Armunk, NY) and Microsoft Excel-2021. All analyses were intention-to-treat.

### Trial Registration

ClinicalTrials.gov NCT05635539. Registered 1 December 2022.

## 3 RESULTS

The symptoms of COVID and Influenza were the common typical such as fever in 62 patients (100%), cough in 48 (77.4%), runny or stuffy nose in 51 (82.2%), fatigue in 49 (79%), chills in 30 (48.4%), sore throat in 14 (22.6%), headache in 22 (35.4%), body aches in 18 (29%). Loss of taste or smell was in 21 patients (33.8%): 9 patients with COVID; 12 patients with Influenza. This symptom did not differ between these diseases (95%; OR 0.96, CI 0.34-2.73, *P*=0.46). Loss of appetite and smell irritability were in 51 patients (82.3%): 24 patients with COVID; 26 patients with Influenza (95%; OR 1.19, CI 0.81-1.77, *P*=0.92). Other clinical symptoms of the diseases did not also differ from each other.

Baseline/ 7-day/ 14-day treatment and to 6-week follow-up results concerning anthropometrical data, body composition, and metabolic data are shown in **Table 1**. Patients with overweight lost from 8 to 11 kg (−12,4% from baseline), and it was higher than patients without overweight (−9,14% from baseline; -9.7±0.7 kg vs. -6.4±0.6 kg, respectively; *P*<0.001) at 14-day of the treatment. In patients with overweight BMI at 14-day was highly significantly decreased (−4.2 kg/m^2^) than in patients without overweight (−2.2 kg/m^2^) (*P*<0.001).

**Table 1.**
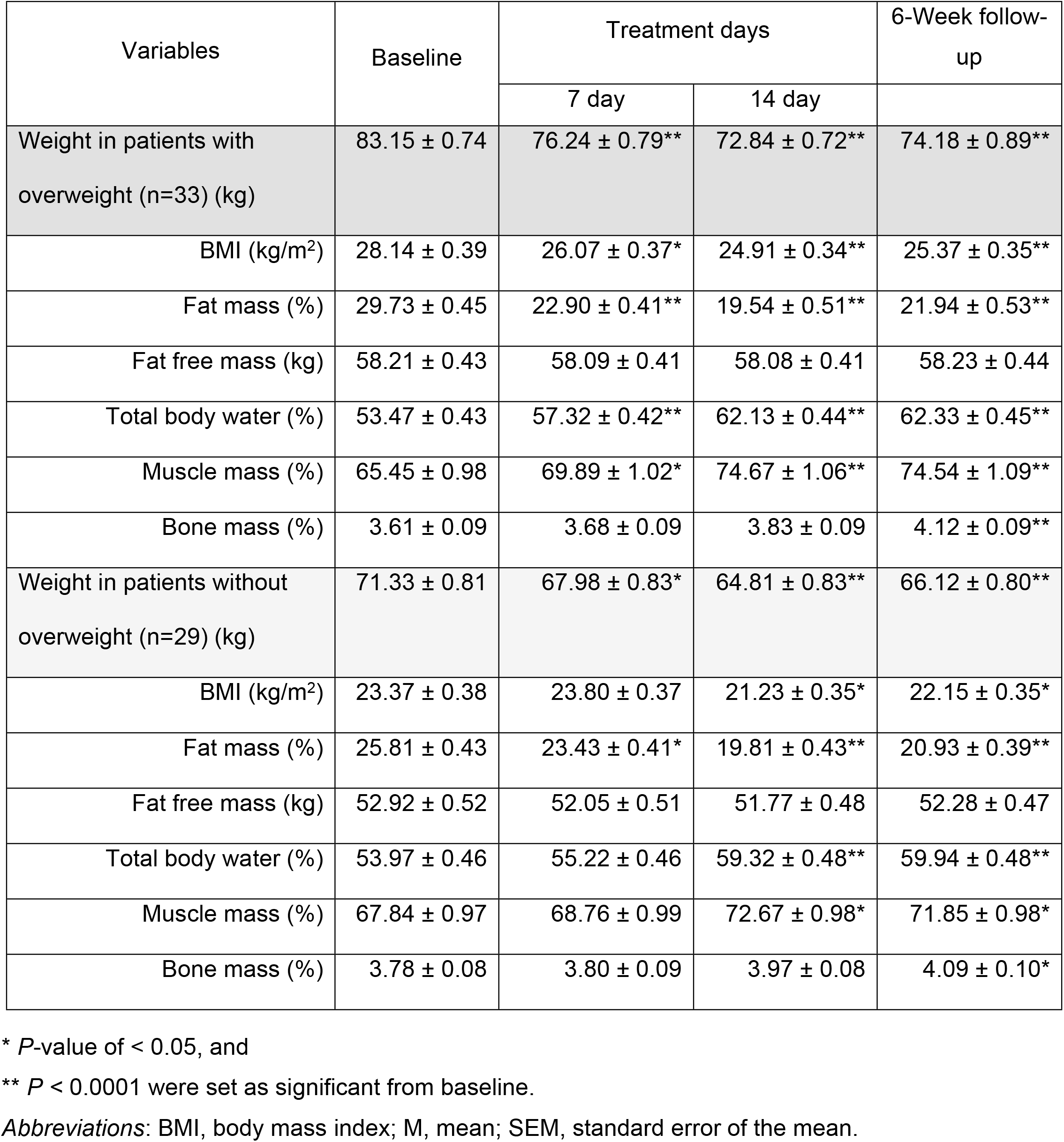
Anthropometrical data, body composition in patients (with/ without overweight) with COVID and Influenza in comorbidity with the Non-communicable diseases before (baseline), during treatment, and 6-week follow-up (M ± SEM)

| Variables | Baseline | Treatment days |  | 6-Week follow-up |
| --- | --- | --- | --- | --- |
|  |  | 7 day | 14 day |  |
| Weight in patients with overweight (n=33) (kg) | 83.15 ± 0.74 | 76.24 ± 0.79** | 72.84 ± 0.72** | 74.18 ± 0.89** |
| BMI (kg/m<sup>2</sup>) | 28.14 ± 0.39 | 26.07 ± 0.37* | 24.91 ± 0.34** | 25.37 ± 0.35** |
| Fat mass (%) | 29.73 ± 0.45 | 22.90 ± 0.41** | 19.54 ± 0.51** | 21.94 ± 0.53** |
| Fat free mass (kg) | 58.21 ± 0.43 | 58.09 ± 0.41 | 58.08 ± 0.41 | 58.23 ± 0.44 |
| Total body water (%) | 53.47 ± 0.43 | 57.32 ± 0.42** | 62.13 ± 0.44** | 62.33 ± 0.45** |
| Muscle mass (%) | 65.45 ± 0.98 | 69.89 ± 1.02* | 74.67 ± 1.06** | 74.54 ± 1.09** |
| Bone mass (%) | 3.61 ± 0.09 | 3.68 ± 0.09 | 3.83 ± 0.09 | 4.12 ± 0.09** |
| Weight in patients without overweight (n=29) (kg) | 71.33 ± 0.81 | 67.98 ± 0.83* | 64.81 ± 0.83** | 66.12 ± 0.80** |
| BMI (kg/m<sup>2</sup>) | 23.37 ± 0.38 | 23.80 ± 0.37 | 21.23 ± 0.35* | 22.15 ± 0.35* |
| Fat mass (%) | 25.81 ± 0.43 | 23.43 ± 0.41* | 19.81 ± 0.43** | 20.93 ± 0.39** |
| Fat free mass (kg) | 52.92 ± 0.52 | 52.05 ± 0.51 | 51.77 ± 0.48 | 52.28 ± 0.47 |
| Total body water (%) | 53.97 ± 0.46 | 55.22 ± 0.46 | 59.32 ± 0.48** | 59.94 ± 0.48** |
| Muscle mass (%) | 67.84 ± 0.97 | 68.76 ± 0.99 | 72.67 ± 0.98* | 71.85 ± 0.98* |
| Bone mass (%) | 3.78 ± 0.08 | 3.80 ± 0.09 | 3.97 ± 0.08 | 4.09 ± 0.10* |

\* *P*-value of < 0.05, and
\*\* *P* < 0.0001 were set as significant from baseline.
*Abbreviations:* BMI, body mass index; M, mean; SEM, standard error of the mean.

Weight loss in both groups was due to reduction of fat mass (*P*<0.0001). Percentages of total body water and muscle mass tended to increase significantly in both groups at 7-day/14-day of the treatment, and the percentage of bone mass increased significantly too (in patients with overweight *P*<0.0001; in patients without overweight *P*<0.05). Lean body mass (fat free mass) did not change significantly during weight loss in both groups, patients with and without overweight (*P* = 0.82-0.97). These trends persisted at 6-week of follow-up.

Starting from 2-3 days of the treatment, in almost patients is increased a sputum production to 1.0-1.5 liters a day. A sputum production decreased by 7-9 days of the treatment.

Body temperature decreased starting from 3-4 days of treatment, and it is normalized to 6-9 days, and also was normal at 6-week follow-up (**Table 2**). Begin from 3-5 days of the treatment, in most of the patients their urine became turbid, muddy and intensively darkly colored, which persisted for several days. Urine microscopy showed organic and non-organic salts such as oxalates, urates, phosphates, and carbonates of calcium and magnesium, and leukocyturia (20-35 per in sight). Starting from 5-7 days of the treatment, the patients noticed a physical relief, increase in physical/mental workability, and exercise tolerance. Infection detection at 14-day of treatment was a negative in 26 patients with COVID of 27, and 34 patients with Influenza of 35. Re-detection on the next two days showed a negative in both patients.

**Table 2.** Body temperature, Blood pressures, Inflammation level, Glucose and Lipids metabolism, Lipid and protein oxidative products, and chest CT in patients with COVID and Influenza in comorbidity with the Non-communicable diseases at baseline, during treatment, and 6-week follow-up (M ± SEM)

| Variables (n) | Baseline | Treatment days |  | 6-Week follow-up |
| --- | --- | --- | --- | --- |
|  |  | 7 day | 14 day |  |
| Body temperature, °C (62) | 39.34 ± 0.08 | 37.86 ± 0.08** | 36.7 ± 0.05** | 36.5 ± 0.03** |
| Systolic BP in patients with hypertension, mmHg (38) | 161.3 ± 1.31 | 116.2 ± 0.77** | 118.3 ± 0.46** | 120.8 ± 1.17** |
| Diastolic BP in patients with hypertension, mmHg (38) | 101.6 ± 0.85 | 78.92 ± 0.67** | 80.89 ± 0.66** | 81.23 ± 0.79** |
| Total protein, g/l (62) | 62.41 ± 0.63 | -- | 66.92 ± 0.65** | 71.43 ± 0.67** |
| Hemoglobin, g/L (62) | 128.6 ± 0.71 | -- | 131.7 ± 0.72* | 136.9 ± 0.78** |
| White blood cells, $\times 10^9$/L (62) | 12.29 ± 0.17 | 11.83 ± 0.16 | 7.68 ± 0.12** | 5.21 ± 0.09** |
| Neutrophil, $\times 10^9$/L (62) | 7.128 ± 0.012 | 6.389 ± 0.010** | 3.610 ± 0.009** | 2.501 ± 0.012** |
| Lymphocytes, $\times 10^9$/L (62) | 2.581 ± 0.008 | 3.256 ± 0.009** | 2.918 ± 0.009** | 1.824 ± 0.008** |
| Neutrophil-to-lymphocyte ratio (62) | 2.762±0.009 | 1.962 ± 0.009** | 1.237 ± 0.009** | 1.371 ± 0.008** |
| Erythrocyte sedimentation rate, mm/Hour (62) | 33.4 ± 0.56 | -- | 13.76 ± 0.24** | 7.59 ± 0.24** |
| Total Fibrinogen, g/L (62) | 4.83 ± 0.07 | 5.25 ± 0.09* | 3.81 ± 0.08** | 2.92 ± 0.09** |
| C-reactive protein, mg/L (62) | 34.86 ± 1.26 | 26.35 ± 1.37** | 12.39 ± 0.79** | 6.57 ± 0.74** |
| Glucose in patients with T2D, mmol/L (26) | 12.86 ± 0.19 | 6.75 ± 0.11** | 4.60 ± 0.09** | 4.58 ± 0.10** |
| HbA1c, % (26) | 7.03 ± 0.09 | -- | -- | 4.86 ± 0.08** |
| Immunoassay Insulin, nU/L (26) | 22.38 ± 0.58 | -- | 9.61 ± 0.22** | 6.45 ± 0.17** |
| HOMA-IR index (26) | 12.79 ± 0.35 | -- | 1.96 ± 0.17** | 1.31 ± 0.17** |
| Platelets, $\times 10^9$/L (51) | 186.5 ± 4.6 | 196.6 ± 4.7 | 238.5 ± 5.8** | 278.3 ± 6.9** |
| ALT, U/L (51) | 134.3 ± 5.4 | 102.6 ± 5.7* | 78.4 ± 4.2** | 27.5 ± 1.8** |
| AST, U/L (51) | 166.5 ± 5.5 | 126.3 ± 5.9** | 92.4 ± 4.9** | 29.3 ± 1.8** |
| Urea, mmol/L (62) | 5.17 ± 0.07 | 7.54 ± 0.07** | 5.62 ± 0.06** | 4.14 ± 0.07** |
| Creatinine, µmol/L (62) | 75.58 ± 1.27 | 109.73 ± 1.59** | 62.86 ± 1.07** | 58.45 ± 0.94** |
| Cholesterol, mmol/L (62) | 5.82 ± 0.07 | 6.83 ± 0.08** | 6.19 ± 0.09* | 4.74 ± 0.07** |
| Triglyceride, mmol/L (62) | 2.52 ± 0.07 | 5.94 ± 0.09** | 5.73 ± 0.08** | 1.11 ± 0.05** |
| HDL, mmol/L (62) | 0.96 ± 0.03 | 1.02 ± 0.03 | 1.06 ± 0.03* | 1.68 ± 0.04** |
| MDA, µmol/L (62) | 60.74 ± 1.84 | -- | 41.51 ± 0.87** | 31.37 ± 0.86** |
| AOPP, µmol/L (62) | 295.6 ± 9.2 | -- | 206.3 ± 11.1** | 188.3 ± 7.4** |
| SOD, U/mg (62) | 34.58 ± 1.07 | -- | 57.72 ± 1.06** | 66.53 ± 0.98** |
| Catalase, U/g (62) | 26.52 ± 0.67 | -- | 46.83 ± 0.65** | 49.29 ± 0.71** |
| Chest CT scan, score (44) | 13.12 ± 0.38 | -- | 1.72 ± 0.12** | 0.0 ± 0.0 |

\* *P*-value of < 0.05, and
\*\* *P* < 0.0001 were set as significant from baseline.
*Abbreviations:* AOPP, Advanced oxidation protein products; BP, blood pressure; CT, Computed tomography; HDL, high density lipoprotein; HOMA-IR, Homeostasis Model Assessment for Insulin Resistance; M, mean; MDA, Malondialdehyde; SOD, Superoxide dismutase; SEM, standard error of the mean; T2D, type 2 diabetes.

The weight loss treatment evoked significantly rising of serum urea and creatinine on 7-day of treatment. We noticed normalization levels of serum urea and creatinine on 14-day. (**Table 2**) During lipolysis on the weight loss treatment there was no protein lost because total serum protein level significantly increased from baseline (62.41±0.53 g/l) to 14-day treatment (66.92±0.65 g/l, *P*<0.0001), and 6-week follow-up (71.43±0.67 g/l, *P*<0.0001). Hemoglobin level also significantly increased from baseline (128.6±0.71 g/l) to 14-day treatment (131.7±0.72 g/l, *P*<0.05), and 6-week follow-up (136.9±0.78 g/l, *P*<0.0001). HDL significantly increased from baseline (0.96±0.03 mmol/L) to 14-day treatment (1.06±0.03 mmol/L, *P*=0.02), and 6-week follow-up (1.68±0.04 mmol/L, *P*<0.0001). (**Table 2**)

White blood cells were gradually decreased on the 14 day. Relative lymphocyte count increased that the NLR was significantly decreased (*P*<0.0001). Total fibrinogen, C-reactive protein, and Erythrocyte sedimentation rate were significantly decreased to normal at 14-day of treatment (*P*<0.0001) and 6-week follow-up (*P*<0.0001). (**Table 2**)

The weight loss treatment led to a normal systolic/diastolic BP starting from 7-day of treatment, and it were normalized at 14-day and 6-week follow-up (**Table 2**) reaching the levels recommended by the American Heart Association (2014).

The positive changes in glucose metabolism in patients with T2D (n=26) were at 14-day of treatment and also at 6-week follow-up. Fasting Glucose and Immunoassay Insulin, and HOMA-IR index quickly decreased at 14-day of treatment. Insulin decreased 2.3-fold from baseline at 14-day of treatment, whereas 3.5-fold from baseline at 6-week follow-up. HbA1c decreased to the normal by 6-week (4,86±0.08) by 31% (*P*<0.05).

Cholesterol and triglycerides are significantly increased by 17.2-6.4% and 68.8-62.7% from baseline at 7 and 14 days of the treatment, respectively.

In patients with NASH (n=51) ALT and AST levels were significantly decreased from baseline (134.3±5.4 and 166.5±5.5 U/L, respectively) at 7-day (by 102.6±5.7 (*P*<0.05) and 126.3±5.9 U/L (*P*<0.0001), respectively); and 14-day (by 78.4±4.2 (*P*<0.0001) and 92.4±4.9 U/L (*P*<0.0001), respectively). On the 6-week follow-up ALT and AST levels returned to normal (*P*<0.0001). (**Table 2**)

Platelets in patients with NASH (n=51) significantly increased from baseline 186.5±4.6, ×10^9^/L at 14-day of treatment by 238.5±5.8, ×10^9^/L (*P*<0.0001), and at 6-week follow-up by 278.3±6.9, ×10^9^/L (*P*<0.0001).

### Abnormal chest imaging

CT-scans were abnormal in 44 patients (70.97%) aged >40 years old, including 8 patients with NASH, T2D and Hypertension; 6 patients with NASH and T2D; 12 patients with T2D and Hypertension; 18 patients with NASH and Hypertension.

CT findings included multiple small patchy shadows, centrilobular nodules and bronchial wall thickening, interstitial inflammation, predominantly distributed in the peripheral of the lungs, ground glass opacities and infiltrates in the lungs, bronchopneumonia patterns. The most common finding was ground glass opacity (n=39; 88.6% including 25 patients with COVID), not followed by consolidation. Pleural effusion and cavitation was not detected. Unilateral lung involvement (n=36; 81.8%) was more common than bilateral involvement (n=6; 13.6%). The right/ left lower zones (n=34; 77.3%) were more commonly affected than the right/ left middle (n=9; 20.5%). Multifocal distribution was more common (n=24, 54.5%) than unifocal distribution (n=19, 43.2%). Chest CT scans were non-specific for the ARDs.

The mean score of chest CT for the patients (n=44) was 13.12±0.38. A significant positive changes was found on 14-day of treatment with the score 1.72±0.12 (*P*<0.0001). There were not found the lung abnormalities at 6-week follow-up (**Table 2**). By 14-day of treatment CT scans showed that the scope of the lesions had been reduced, the density was gradually decreased, the number of lesions decreased, and the ground glass opacities were absorbed. There was no the 6-week risk of arterial thromboembolism and venous thromboembolism among the patients with COVID or Influenza.

The oxidative products of lipids and proteins had a trend to normalize at 14-day treatment and 6-week follow-up, where MDA decreased by 31.7% at 14-day (*P*<0.0001) and by 48.4% at 6-week from baseline (*P*<0.0001), AOPP decreased by 30.2% at 14-day (*P*<0.0001) and by 36.3% at 6-week from baseline (*P*<0.0001), and SOD increased by 66.9% (*P*<0.0001) and Catalase by 76.6% at 14-day (*P*<0.0001), and by 92.4% and 85.8%, respectively, at 6-week from baseline (*P*<0.0001) (**Table 2**).

As the clinical status of the patients was improving, the previous antidiabetic (patients with T2D, n=26), antihypertensive (patients with hypertension, n=38), anti-inflammatory and hepatoprotective (patients with NASH, n=51), and other symptomatic conventional medications were adequately decreased starting from 2-5 days of ANADETO treatment. By days of 5 to 8 after the treatment start, the drugs were stopped completely. There were no recurrence of T2D, hypertension, and NASH at 6-week follow-up.

## 4. DISCUSSION

Our data showed that the fast weight loss ‘ANADETO’ method (−12,4% fat loss in patients with overweight, and -9,14% fat loss in patients without overweight) at 14-day treatment and at 6-week follow-up had a clinical, laboratory and imaging effectiveness in patients with COVID and Influenza in comorbidity with T2D, hypertension, and NASH.

The weight loss in patients with overweight goes faster than in patients without overweight. This an interesting finding in the physiology of weight loss might be explained by different structure of fat in people with and without overweight. (49) People who quickly weight gain can accumulate a lightweight body fat, and people who slowly weight gain can accumulate a dense body fat. Some people cannot weight gain that possibly related with genotype, different fat structure, or metabolic equivalent when the same amount of fat can give different calorie. (50, 51)

The weight loss improved an immunity of the patients by significantly increased in lymphocyte count and decreased in NLR. In fact, lymphopenia has also been associated with severe illness and poorer survival in COVID-19. (52, 53) In clinical practice, it was observed that in some non-survivors, the absolute value of lymphocytes decreases progressively while the white blood cell count gradually increases over time, resulting in a “divergence” between the absolute value of neutrophils and lymphocytes, that NLR, may be correlated with the progression and prognosis of this newly emerged disease. (54)

Our clinical study confirmed that weight loss significantly reduces inflammation biomarkers. (55, 56) Reducing inflammation through weight loss could be associated with reduced risk for cardiovascular disease and other obesity-associated chronic diseases. (28, 57) The weight loss has immunomodulatory effect, repealed insulin resistance, improved blood insulin, increased blood hemoglobin, and improved blood pressure. (58, 59) The weight loss method also decreased blood cholesterol/triglyceride, lipid/ protein oxidative products, and increased anti-oxidative enzymes.

Increased serum urea and creatinine during 14-day weight loss can testify about endogen intoxication related to active lipolysis. Adsorbed endogen organic toxins in adipocytes are eliminated through blood system during lipolysis. (60, 61) Increase in lipids (cholesterol and triglyceride) from baseline at 7 and 14 days of the treatment confirms the activity of lipolysis during the weight loss treatment.

More sodium ions are eliminated through urine in patients with acute respiratory inflammation in comorbidity with NCDs. (62, 63) Blood and urine sodium became normal on a weight loss due to elimination of metabolic pollutants from the body.

Improvements of liver function we noticed by decrease in ALT and AST levels and increase in blood platelets, and serum total protein. Weight loss improves liver health, cardiovascular risk, and quality of life, therefore some authors offer it as feasible treatment option for patients with NASH. (64, 65)

The study evidenced that chest CT in the patients with COVID and Influenza are highly variable and nonspecific, that is considered with other authors. (66) Chest CT has been widely used during the COVID pandemic, which has a high sensitivity, about 60-97%, but low specificity (about 20-50%). (66, 67) Many radiology professional societies recommend against performing chest CT as a primary technique for the diagnosis of COVID pneumonia. (68) In patients with ARDs, CT findings should be interpreted in combination with a clinical context.

Overweight is a biological burden for the body, which consumes additional immunological, antitoxic, trophic, excretion function. (69) The ANADETO method metabolizes ‘old lipids’ with fat loss, decreases in inflammation, increases in immune response, improves of glucose and lipid metabolism, and liver functions. The patients experienced side effects related to symptoms of metabolic intoxication that is a common problem during weight loss. Adipose tissue absorbs different persistent organic pollutants, (70) and plays an important role in storing of different pollutants. (71)

The ability to accumulate adipose tissue is one of the most important adaptive mechanisms for survival, but now we observe a steady increase in obesity and related diseases. (72, 73)

The fast weight loss method was a safe, well tolerated, and acceptable nutritional therapy option for patients with acute respiratory inflammation diseases in comorbidity with T2D, hypertension, and NASH. The optimal mix of caloric restriction, sodium intake, walking (at normal body temperature), and sexual self-restraint behavior would be an effective treatment method of ARDs (COVID and Influenza) in comorbidity with NCDs.

## 5. CONCLUSIONS

Thus, the fast weight loss ‘ANADETO’ cured clinical, laboratory, and instrumental data of inflammation, improved glucose and lipids metabolism, normalized systolic/diastolic BPs and NASH’s biochemical outcomes, decreased reactive oxygen species, and allowed drugs reduction up to its complete withdrawal in patients with COVID and Influenza in comorbidity with T2D, hypertension, and NASH. The weight loss occurred only due to a decrease in fat mass.

### Study limitation

The study has several limitations. First, the study was short duration. Second, we did not study T-cell and B-cell subpopulation. Third, the clinical trial had approximately 13.9% of the assigned population dropped out prior to completion. Fourth, the study was non-randomized, non-controlled trial. Fifth, published studies about positive role of weight loss in patients with COVID and Influenza in comorbidity with Non-communicable diseases are limited in scope and number. Further randomized controlled trials with longer-term follow-up are needed to confirm and extend the results of the study.

## Data Availability

First, the data is too large (> 2.8 GB). Secondly, we will make our data available to any investigator/reviewer on their own request, so that the personal privacy of our patients cannot be compromised.

## List of Abbreviations

ALT: alanine aminotransferase
ANADETO: ‘Analimentary detoxication’ weight loss method
AOPP: advanced oxidation protein products
AST: aspartate aminotransferase
BMI: body mass index
BP: blood pressure
CT: Computed tomography
COVID: SARS-CoV-2, a virus which causes the disease known as COVID-19
HbA1c: glycosylated hemoglobin A1c
HDL: high density lipoprotein
HOMA-IR: Homeostasis Model Assessment for Insulin Resistance
M±SD: Mean ± standard deviation
M±SEM: Mean ± standard error of the mean
MDA: malondialdehyde
NASH: Nonalcoholic steatohepatitis
NCDs: non-communicable chronic diseases
ROS: reactive oxygen species
SOD: superoxide dismutase
T2D: type 2 diabetes mellitus

## DECLARATIONS

### Ethics approval and consent to participate

The Local Ethics Committee of the University Medical Center (phone: +71272-692586; https://umc.org.kz/en/?ethics-commission) approved a study “The fast weight loss in patients with acute and chronic respiratory infections in comorbidity with non-communicable diseases” (approval protocol number is #7 of 26.09.2019).

### Consent for publication

All authors of the manuscript affirm that they had access to the study data and reviewed and approved the final version.

### Author contributions

*KO:* study design, collection of the clinical data, diagnosis and treatment of the patients, bibliography review, statistical analysis, data interpretation, writing of the paper. *AD:* study design, data collection, diagnosis and treatment of the patients, data interpretation, bibliography and paper review. *AI:* study design, data collection, writing the Introduction and Methods, data interpretation, paper review. *ZZ* and *MG:* preparation of the statistical data in Excel, collection of the clinical data, bibliography and paper review, data interpretation, statistical analysis, writing of the Methods and Discussion, paper review. *GB:* study design, data collection, writing the results and discussion, and paper review. *AT:* writing of the discussion, statistical analysis, paper review. *AN:* collection of the clinical data, preparation of statistical data in Excel, patient diagnosis, bibliography search and review. *BD:* study design, data collection and interpretation, writing the discussion, bibliography and paper review.

All authors read and approved the final manuscript.

## ACKNOWLEDGMENTS

The authors thank the Republican Diagnostic Center at University Medical Center and ANADETO medical center in recruiting patients and collecting data for the study, and technical assistance.

